# The UGT2A1/UGT2A2 locus is associated with COVID-19-related anosmia

**DOI:** 10.1101/2021.05.28.21257993

**Authors:** Janie F. Shelton, Anjali J. Shastri, The 23andMe COVID-19 Team, Stella Aslibekyan, Adam Auton

## Abstract

Loss of sense of smell is a characteristic symptom of infection with SARS-CoV-2. However, specific mechanisms linking infection with loss of smell are poorly understood. Using self-reported symptom data from the 23andMe COVID-19 study, we describe the demographic patterns associated with COVID-19 related anosmia, and find the symptom is more often reported in women and younger respondents, and less often by those of East Asian and African American ancestry compared to those of European ancestry. We ran a trans-ethnic genome-wide association study (GWAS) comparing loss of smell or taste (n=47,298) with no loss of smell or taste (n=22,543) among those with a positive SARS-CoV-2 test result. We identified an association (rs7688383) in the vicinity of the *UGT2A1 and UGT2A2* genes (OR=1.115, p-value=4×10^−15^), which have been linked to olfactory function. These results may shed light on the biological mechanisms underlying COVID-19 related anosmia.

## Introduction

Loss of sense of smell, or anosmia, is the main neurological symptom and one of the earliest and most commonly reported indicators of the acute phase of SARS-CoV-2 infection. While a large fraction of COVID-19 patients report loss of smell or taste, the underlying mechanism remains unclear.^1^

23andMe is a direct-to-consumer genetic testing company with over 10 million genotyped customers. As part of the 23andMe service, customers are genotyped on SNP microarrays and offered the opportunity to participate in scientific research, and approximately 80% of customers consent to do so. In general, research participation is conducted via online surveys, which research participants can complete at any time. Research participants are re-contactable and can be invited to participate in new surveys that are developed over time.

Here, we conducted a genome-wide association study (GWAS) of COVID-19-related anosmia, having collected data from over one million research participants as previously described.^2^ By asking 23andMe research participants to report the symptoms they encountered during their COVID-19 experience, we identified 47,298 individuals who reported loss of taste or smell along with a SARS-CoV-2 positive test, and 22,543 individuals that reported a positive test but did not experience loss of taste or smell. We performed GWAS separately in samples of European, Latino, African American, East Asian, and South Asian ancestries and used the resulting data to perform a trans-ethnic meta-analysis.

We identified a strong association at a locus containing the *UGT2A1* and *UGT2A2* genes. These genes, part of a family of uridine diphosphate glycosyltransferases, are expressed in the olfactory epithelium and function as odorant metabolizing enzymes, facilitating the elimination of exogenous and endogenous compounds and playing a critical role in the physiology of olfaction.^3^ Recent evidence suggests that SARS-CoV-2 impedes olfactory sensation by infecting and compromising the essential functions of olfactory support cells,^4^ which may be modulated by *UGT2A1/UGT2A2*. Our findings represent the first genetic link to the underlying mechanisms of COVID-19-related anosmia.

## Methods

### Overview of study recruitment and data collection

Participants in this study were recruited from the customer base of 23andMe, Inc., a personal genetics company. Participants provided informed consent and participated in the research online, under a protocol approved by the external AAHRPP-accredited IRB, Ethical & Independent Review Services (E&I Review). Participants were included in the analysis on the basis of consent status as checked at the time data analyses were initiated.

Full details of the data collection paradigm for this study have been described previously.^2^ In brief, primary recruitment was carried out by email to approximately 6.7 million 23andMe research participants over 18 years of age and living in the United States or the United Kingdom. Additionally, pre-existing customers were invited to participate in the study through promotional materials on the 23andMe website, the 23andMe mobile application, and via social media. Study participation consisted solely of web-based surveys, including an initial baseline survey, and three follow-up surveys fielded 1-month following completion of the baseline survey. Enrollment continued after the initial recruitment efforts until a data-freeze was taken for this study in March 2021, when 1.3 million participants had completed the baseline survey.

### Phenotype definitions for GWAS

Using the information derived from the surveys, we defined a single phenotype to contrast COVID-19-positive individuals that experienced COVID-19-related anosmia from those that did not. Specifically, participants were asked to respond to the question; “Have you been tested for COVID-19 (coronavirus)?”, with possible responses; “Yes, it was positive / Yes, it was negative / No / My results are pending / I’m not sure”. Of those that responded “Yes, it was positive”, we further considered the question; “During your illness, did you experience any of the following symptoms?” to which participants could select as many as needed from the following list of responses, “Muscle or body aches / Fatigue / Dry cough / Sore throat / Coughing up of sputum or phlegm (productive cough) / Loss of smell or taste / Chills / Difficulty breathing or shortness of breath / Pressure or tightness in upper chest / Diarrhea / Nausea or vomiting / Sneezing / Loss of appetite / Runny nose / Headache / Intensely red or watery eyes”. We defined cases as SARS-CoV-2 test positive individuals that also reported “Loss of smell or taste”, and controls as SARS-CoV-2 test positive individuals who did not report “Loss of smell or taste”. While some participants reported a COVID-19 diagnosis absent a confirmed positive test for SARS-CoV-2, we did not include such individuals within this analysis.

### Descriptive statistics

Sample sizes and proportions were calculated by age, sex, and ancestry. Differences in anosmia by sex were statistically evaluated with a chi-square statistics, and mean difference in age were evaluated with a t-test. A logistic regression model was constructed to evaluate anosmia as a function of ancestry, age (categorical), and sex. All analyses were conducted in R statistical software version 3.6.3.

### Genotyping and SNP imputation

DNA extraction and genotyping were performed on saliva samples by CLIA-certified and CAP-accredited clinical laboratories of Laboratory Corporation of America. Samples were genotyped on one of five genotyping platforms. The V1 and V2 platforms were variants of the Illumina HumanHap550 + BeadChip and contained a total of about 560,000 SNPs, including about 25,000 custom SNPs selected by 23andMe. The V3 platform was based on the Illumina OmniExpress + BeadChip and contained a total of about 950,000 SNPs and custom content to improve the overlap with our V2 array. The V4 platform was a fully custom array of about 950,000 SNPs and included a lower redundancy subset of V2 and V3 SNPs with additional coverage of lower-frequency coding variation. The V5 platform was based on the Illumina GSA array, consisting of approximately 654,000 pre-selected SNPs and approximately 50,000 custom content variants. Samples that failed to reach 98.5% call rate were re-analyzed. Individuals whose analyses failed repeatedly were re-contacted by 23andMe customer service to provide additional samples, as is done for all 23andMe customers.

Participant genotype data were imputed using the Haplotype Reference Consortium (HRC) panel^5^, augmented by the Phase 3 1000 Genomes Project panel^6^ for variants not present in HRC. We phased and imputed data for each genotyping platform separately. For the non-pseudoautosomal region of the X chromosome, males and females were phased together in segments, treating the males as already phased; the pseudoautosomal regions were phased separately. We then imputed males and females together, treating males as homozygous pseudo-diploids for the non-pseudoautosomal region.

### Genome-wide association study (GWAS)

Genotyped participants were included in GWAS analyses on the basis of ancestry as determined by a genetic ancestry classification algorithm.^7^ For each phenotype, we selected a set of unrelated individuals so that no two individuals shared more than 700cM of DNA identical by descent (IBD). For case-control phenotypes, if a case and a control were identified as having at least 700cM of DNA IBD, we preferentially discarded the control from the sample.

For case-control comparisons, we tested for association using logistic regression, assuming additive allelic effects. For tests using imputed data, we use the imputed dosages rather than best-guess genotypes. We included covariates for age, age squared, sex, a sex:age interaction, the top ten principal components to account for residual population structure, and dummy variables to account for genotyping platform. The association test p-value was computed using a likelihood ratio test, which in our experience is better behaved than a Wald test on the regression coefficient. Results for the X chromosome were computed similarly, with men coded as if they were homozygous diploid for the observed allele.

We ran GWAS for each phenotype separately, and combined both genotyped and imputed data. When choosing between imputed and genotyped GWAS results, we favored the imputed result, unless the imputed variant was unavailable or failed quality control (QC). For imputed variants, we removed variants with low imputation quality (r^2^ < 0.5 averaged across batches, or a minimum r^2^ < 0.3) or with evidence of batch effects (ANOVA F-test across batches, p-value < 10^−50^). For genotyped variants, we removed variants only present on our V1 or V2 arrays (due to small sample size) that failed a Mendelian transmission test in trios (p-value < 10^−20^), that failed a Hardy-Weinberg test in Europeans (p-value < 10^−20^), failed a batch effect test (ANOVA p-value < 10^−50^), or had a call rate < 90%.

We repeated the GWAS analysis separately in each population cohort for which we had sufficient data (European, Latino, African American, East Asian, and South Asian), and the resulting summary statistics were corrected for inflation using genomic control when the inflation factor was estimated to be greater than 1. We then performed trans-ethnic meta-analysis using a fixed effects model (inverse variance method^8^), restricting to variants of at least 1% minor allele frequency in the pooled sample, and minor allele count > 30 within each subpopulation. Both the input GWAS and resulting meta-analysis were corrected for inflation using genomic control where necessary.

Within each GWAS, we identified regions with genome-wide significant (GWS) associations. We define the region boundaries by identifying all SNPs with p-value < 10^−5^ within the vicinity of a GWS association, and then grouping these regions into intervals so that no two regions are separated by less than 250 kb. We consider the SNP with the smallest p-value within each interval to be the index SNP. Within each region, we calculated a credible set using the method of Maller *et al*.^9^

## Results

### Respondent characteristics

We previously reported the use of the 23andMe research platform to collect data regarding participants’ experiences with COVID-19 during the pandemic.^2^ From this data, 68% of COVID-19 cases reported loss of smell or taste (47,298 with anosmia out of a total of 69,841 of respondents with a positive SARS-CoV-2 test). Female respondents were more likely than males (72% vs. 61%) to report anosmia (chi-square test, p-value < 2.2×10-16), as were younger individuals (mean age of those with loss of smell = 41 years vs. those without = 45 years, p-value < 2.2×10-16, Welch’s t-test). Among genetically determined ancestral groups, rates of anosmia varied between 63% - 70% (Table 1). As expected, compared to other symptoms surveyed, anosmia was much more common among those with a SARS-CoV-2 positive test compared to those with other cold or flu-like symptoms who tested negative for SARS-CoV-2 (Supplementary Figure 1). In a logistic regression model predicting loss of smell by ancestry, adjusted for age and sex, individuals of East Asian or African American ancestry were less likely to report loss of smell and taste (OR = 0.8 and 0.88 respectively) relative to Europeans (Table 2).

**Table 1:** Sample sizes and percentages comparing self-reported anosmia vs. no anosmia among those with a positive SARS-CoV-2 test result.

|  |  | Positive SARS-CoV-2 test result |  |  |  |  |
| --- | --- | --- | --- | --- | --- | --- |
|  |  | Anosmia | % | No anosmia | % | Total |
|  | Total | 47,298 | 68% | 22,543 | 32% | 69,841 |
| Sex | Female | 31,608 | 72% | 12,562 | 28% | 44,170 |
|  | Male | 15,690 | 61% | 9,981 | 39% | 25,671 |
| Age | < =25 | 6,276 | 71% | 2,620 | 29% | 8,896 |
|  | 26-35 | 13,855 | 73% | 5,134 | 27% | 18,989 |
|  | 36-45 | 10,539 | 70% | 4,552 | 30% | 15,091 |
|  | 46-55 | 8,321 | 67% | 4,080 | 33% | 12,401 |
|  | 56-65 | 5,673 | 62% | 3,522 | 38% | 9,195 |
|  | 66-75 | 2,059 | 51% | 1,945 | 49% | 4,004 |
|  | > 75 | 574 | 45% | 689 | 55% | 1,263 |
| Ancestry | European | 33,336 | 67% | 16,257 | 33% | 49,593 |
|  | Latino | 9,233 | 70% | 3,944 | 30% | 13,177 |
|  | African American or black | 1,860 | 66% | 949 | 34% | 2,809 |
|  | East Asian | 626 | 65% | 338 | 35% | 964 |
|  | South Asian | 284 | 63% | 167 | 37% | 451 |
|  | Other | 1,959 | 69% | 888 | 31% | 2,847 |

**Table 2:** Logistic regression model of age, sex, and ancestry, predicting loss of smell or taste among those with a SARS-CoV-2 positive test.

|  | Model results |  |  |
| --- | --- | --- | --- |
|  | Odds Ratio | 95% confidence interval | p-value |
| European | 1 (REF) | NA | NA |
| Latino | 1.05 | 1.003 - 1.09 | 0.033 |
| African American or Black | 0.88 | 0.81 - 0.96 | 0.003 |
| East Asian | 0.80 | 0.70 - 0.92 | 0.002 |
| South Asian | 0.84 | 0.70 - 1.03 | 0.09 |
| Other | 1.01 | 0.93 - 1.11 | 0.79 |
| Female sex | 1.56 | 1.51 - 1.61 | < 0.0001 |
| Age < 25 | 0.99 | 0.94 - 1.06 | 0.97 |
| 26-35 | 1.14 | 1.09 - 1.20 | < 0.0001 |
| 36-45 | 1.0 (REF) | NA | NA |
| 46-55 | 0.87 | 0.83 - 0.92 | < 0.0001 |
| 56-65 | 0.69 | 0.65 - 0.73 | < 0.0001 |
| 66-75 | 0.47 | 0.43 - 0.50 | < 0.0001 |
| 75 and older | 0.37 | 0.33 - 0.42 | < 0.0001 |

### GWAS

We conducted GWAS within each population separately before performing a trans-ethnic meta-analysis using a fixed-effects model. Each input GWAS was corrected for inflation via genomic control (lambda = 1.029, 1.037, 1.024, 1.042, and 1.071 within the European, Latino, African American, East Asian, and South Asian GWAS respectively), as was the subsequent meta-analysis (lambda = 1.001).

Within the trans-ethnic meta-analysis, we identified a single associated loci in the vicinity of the *UGT2A1* and *UGT2A2* genes on chromosome 4 (chr4q13.3) (Figures 1 and 2). No other locus achieved genome-wide significance in the trans-ethnic meta-analysis or in any of the input populations (Supplementary Figures 2-6). The index SNP at this locus was rs7688383 (C/T, with T being the risk allele. p-value = 1.4e-14, OR = 1.11). This SNP has frequency between 19% and 37%, depending on population (Table 3). While the majority of support within the trans-ethnic analysis comes from the European population (for which we have the largest sample size; Supplementary Figures 7-11), effect sizes are consistent across populations (Figure 3).

**Table 3:** Genome-wide association statistics (odds ratios and p-values) for rs7688383 for each ethnicity, including allele frequencies and sample sizes for each GWAS and the trans-ethnic meta-analysis summary statistics.

| Population | Case / Control N | Allele frequency | Alleles | Odds ratio | 95% CI | p-value |
| --- | --- | --- | --- | --- | --- | --- |
| Europe | 27,786 / 13,903 | 36.8% | C/T | 1.11 | [1.08, 1.15] | 1.78E-11 |
| South Asian | 230 / 143 | 32.2% | C/T | 1.26 | [0.88, 1.82] | 0.206 |
| African American | 1,556 / 804 | 31.4% | C/T | 1.13 | [0.98, 1.29] | 0.0895 |
| Latino | 7,841 / 3,419 | 30.1% | C/T | 1.12 | [1.05, 1.19] | 0.0007 |
| East Asian | 557 / 314 | 19.2% | C/T | 1.18 | [0.91, 1.53] | 0.213 |
| **Meta analysis** | **37,790 / 18,583** |  | **C/T** | **1.12** | **[1.09, 1.15]** | **4.17E-15** |

**Figure 1:**
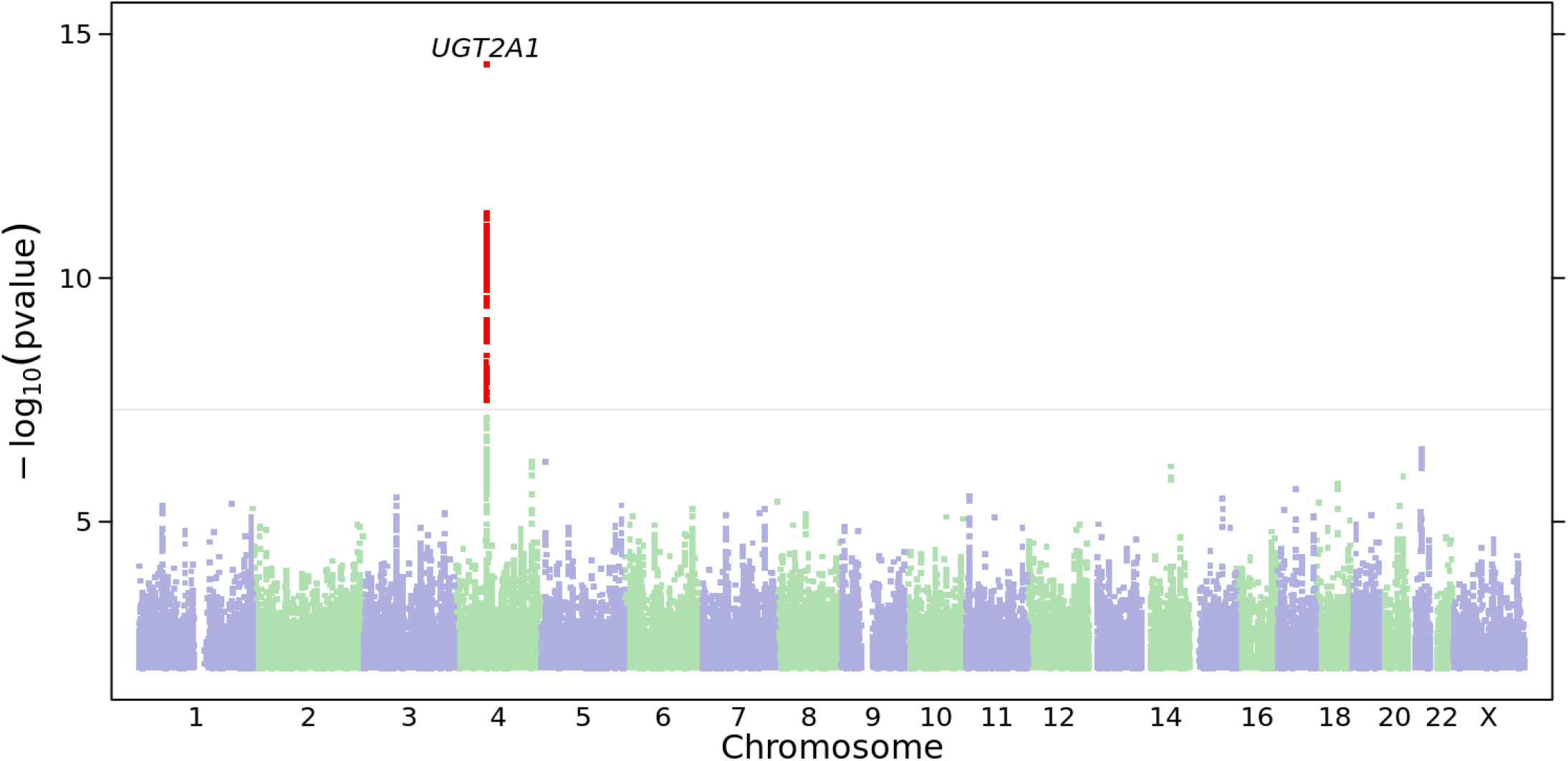
Manhattan plot for the ‘Loss of taste or smell’ phenotype from the trans-ethnic meta analysis. SNPs achieving genome-wide significance are highlighted in red. The nearest gene to the index SNP is indicated above the relevant association peak.

**Figure 2:**
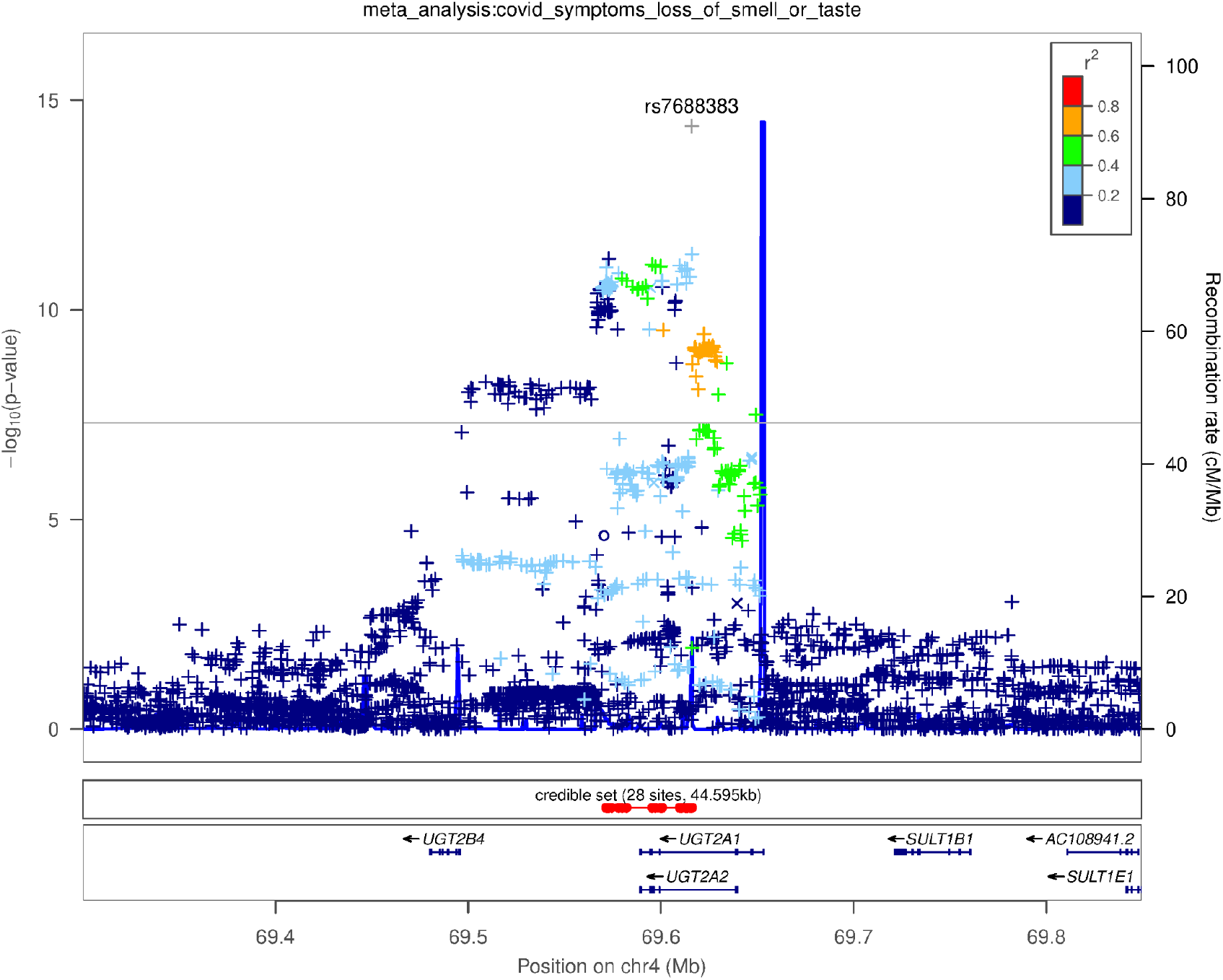
Genetic association with the ‘Loss of taste or smell’ phenotype: regional plot around the *UGT2A1*/*UGT2A2* locus. Colors indicate strength of linkage disequilibrium relative to the index SNP (rs7688383). Imputed variants are indicated with ‘+’ symbols or ‘x’ symbols for coding variants. Where imputed variants weren’t available, directly genotyped variants are indicated by ‘o’ symbols or diamond symbols for coding variants.

**Figure 3:**
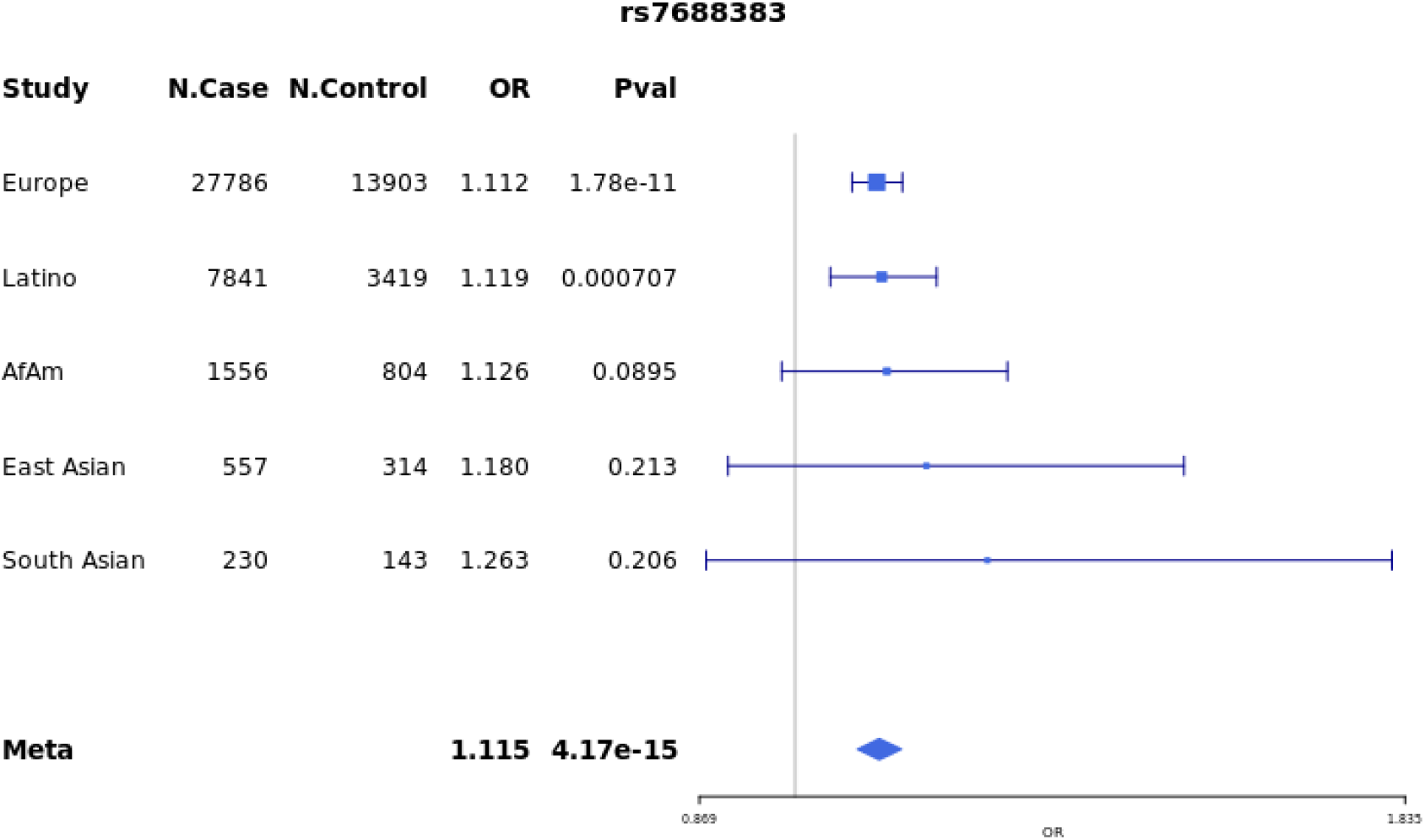
Effect sizes for the index SNP (rs7688383) in each subpopulation and the corresponding meta-analysis.

## Discussion

Loss of smell or taste is a notable symptom of COVID-19. It is distinct from other viral symptoms in its sudden onset^10^, and can occur in the absence of other indications of illness. Here, we report 68% of those with a SARS-CoV-2 test experienced anosmia, which is higher than many other published estimates.^11^ This may be due to the relatively large fraction of milder cases in our study sample compared to clinically ascertained samples, as less severe cases of COVID-19 have been found to more often report anosmia than critical cases.^12^ Further, because the prevalence of this symptom was found to decline with age and is more often reported by females than males, our study sample would be expected to have higher rates of anosmia compared to a case population followed-up after hospitalization.

While many mechanistic explanations have been proposed ^4^ for the COVID-19-related loss of smell, experimental studies suggest that the loss of smell is related to damage to the cilia and olfactory epithelium, but not infection of the olfactory neurons. For example, in an experiment where hamsters were nasally infected with SARS-CoV-2, the olfactory epithelium and cilia became very damaged, which can completely inhibit the ability to smell, but no infection was observed in the olfactory neurons.^13^ Recent evidence suggests that SARS-CoV-2 enters and accumulates in olfactory support cells, specifically, sustentacular cells, which abundantly express the viral entry proteins ACE2 and TMPRSS2 unlike olfactory neurons.^14,15^ These support cells are metabolically and functionally associated with olfactory neurons and with odorant signal transduction (processing odorants by endocytosing the odorant-binding protein complex, detoxifying, maintaining the cilia of mature olfactory receptor neurons, and maintaining epithelial integrity). It has been proposed that olfactory sensation is impaired when these essential functions are disrupted, causing cilial impairment.^4^

We have identified a genetic locus containing two genes (*UGT2A1* and *UGT2A2*) expected to have a relationship with olfactory function. Given their localization and essential function in the metabolization and detoxification of such compounds, these genes may play a role in the physiology of infected cells and the resulting functional impairment that contributes to loss of ability to smell. We hope that the identification of this genetic association with COVID-19 related anosmia may serve as a clue as to how the virus affects cells in the nasal pathway.

## Data Availability

The full set of de-identified summary statistics can be made available to qualified investigators who enter into an agreement with 23andMe that protects participant confidentiality. Interested investigators should visit the following: https://research.23andme.com/covid19-dataset-access/.

## Acknowledgements

We thank the 23andMe research participants and employees who made this study possible. The 23andMe Research Team is:

Aaron A. Petrakovitz, Aaron Kleinman, Adam Auton, Alejandro Hernandez, Anjali J. Shastri, Barry Hicks, Briana Cameron, Catherine H. Weldon, Christophe Toukam Tchakouté, Corinna Wong, Daniella Coker, David A. Hinds, Devika Dhamija, Elizabeth Babalola, Elizabeth S. Noblin, Emily Bullis, Ethan M. Jewett, G. David Poznik, Gabriel Cuellar Partida, Janie F. Shelton, Jared O’Connell, Jessica Bielenberg, Jey McCreight, Jingchunzi Shi, Joanna L. Mountain, Joyce Y. Tung, Karl Heilbron, Katarzyna Bryc, Katelyn Kukar, Keng-Han Lin, Kipper Fletez-Brant, Matthew H. McIntyre, Maya Lowe, Meghan E. Moreno, Morgan Schumacher, Peter Wilton, Pierre Fontanillas, Pooja M. Gandhi, Priyanka Nandakumar, Robert K. Bell, Sarah L. Elson, Sayantan Das, Stella Aslibekyan, Steven J. Micheletti, Suyash Shringarpure, Teresa Filshtein, Vinh Tran, Wei Wang, Will Freyman, Xin Wang, Yunxuan Jiang

## 23andMe COVID-19 Team

Members of the 23andMe COVID-19 Team are: Adam Auton, Adrian Chubb, Alison Fitch, Alison Kung, Amanda Altman, Andy Kill, Anjali Shastri, Antony Symons, Catherine Weldon, Chelsea Ye, Daniella Coker, Janie F. Shelton, Jason Tan, Jeff Pollard, Jennifer McCreight, Jess Bielenberg, John Matthews, Johnny Lee, Lindsey Tran, Maya Lowe, Michelle Agee, Monica Royce, Nate Tang, Pooja Gandhi, Raffaello d’Amore, Ruth Tennen, Scott Dvorak, Scott Hadly, Stella Aslibekyan, Sungmin Park, Taylor Morrow, Teresa Filshtein Sonmez, Trung Le, and Yiwen Zheng. All team members are employees of 23andMe Inc.

## Author Contributions Statement

JFS, AJS, SA, and AA designed this study. The 23andMe COVID-19 Team developed the recruitment and participant engagement strategy and acquired and processed the data. JFS, SA, and AA analyzed the data.

JFS, AJS, and AA interpreted the data. JFS, AJS, and AA wrote the manuscript. All authors participated in the preparation of the manuscript by reading and commenting on drafts prior to submission.

## Tables and Figures

**Supplementary Figure 1:**
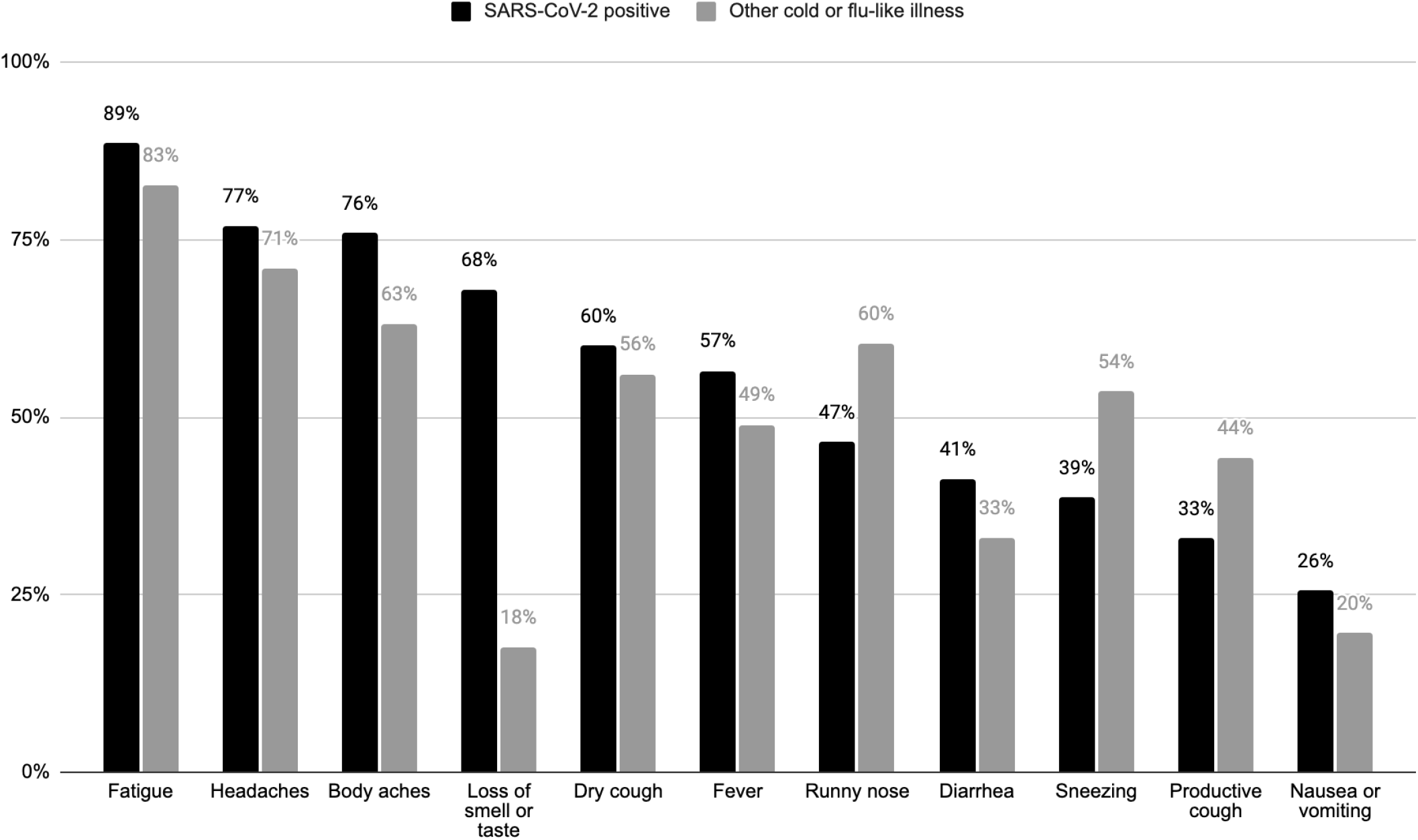
Self-reported symptoms experienced during SARS-CoV-2 infection with a positive test as compared to other cold or flu-like illness accompanying a negative SARS-CoV-2 test.

**Supplementary Figure 2:**
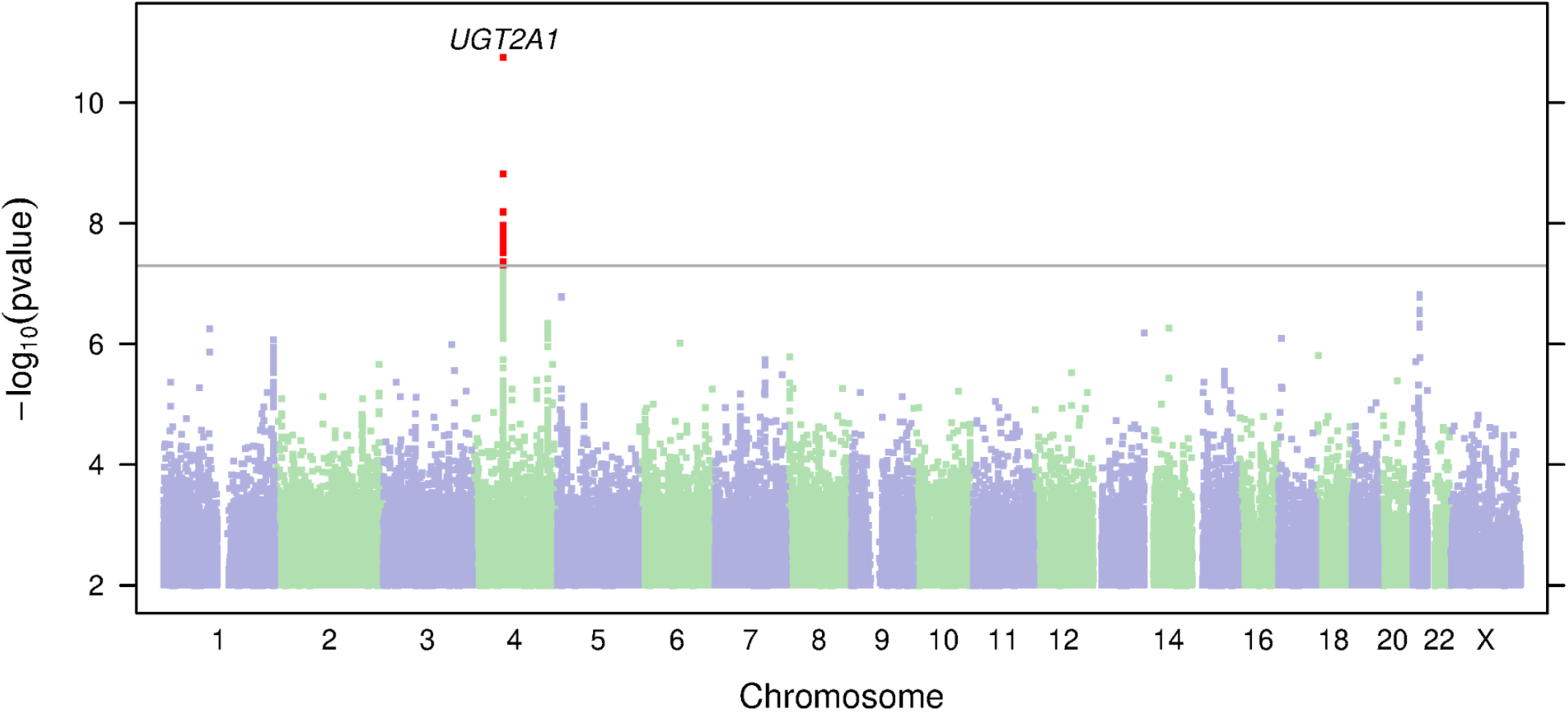
Manhattan plot for the ‘Loss of taste or smell’ phenotype in the European population. SNPs achieving genome-wide significance are highlighted in red. The nearest gene to each index SNP is indicated above the relevant association peaks.

**Supplementary Figure 3:**
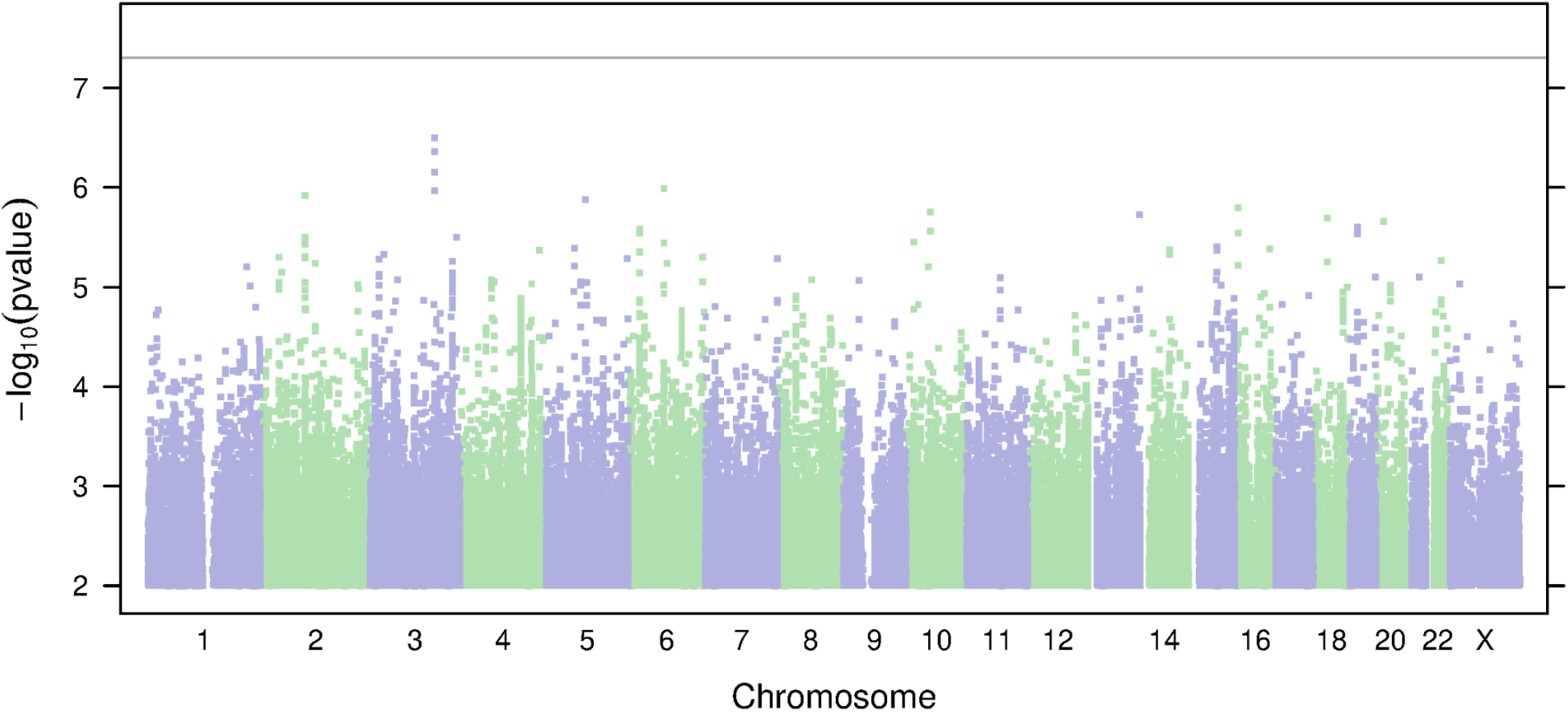
Manhattan plot for the ‘Loss of taste or smell’ phenotype in the Latino population.

**Supplementary Figure 4:**
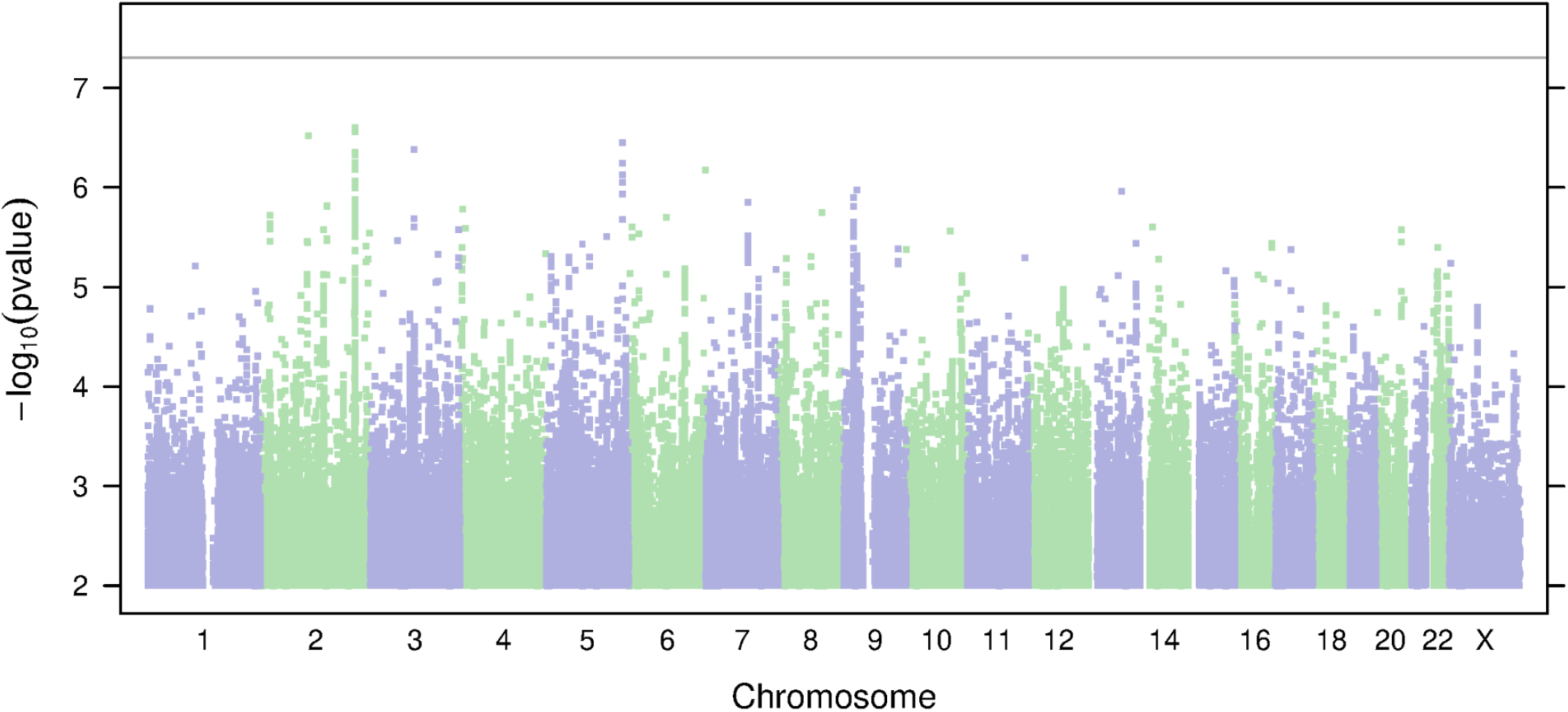
Manhattan plot for the ‘Loss of taste or smell’ phenotype in the African American population.

**Supplementary Figure 5:**
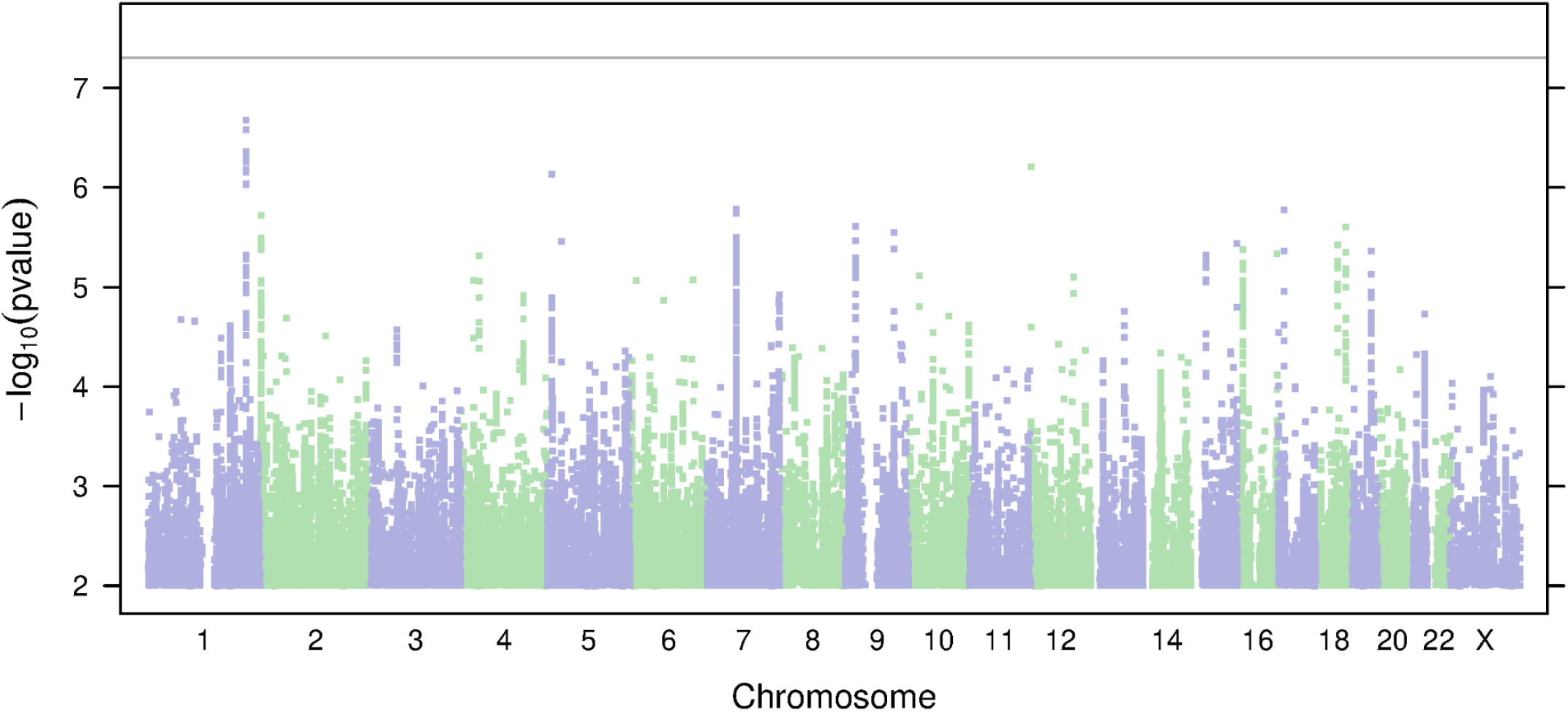
Manhattan plot for the ‘Loss of taste or smell’ phenotype in the East Asian population.

**Supplementary Figure 6:**
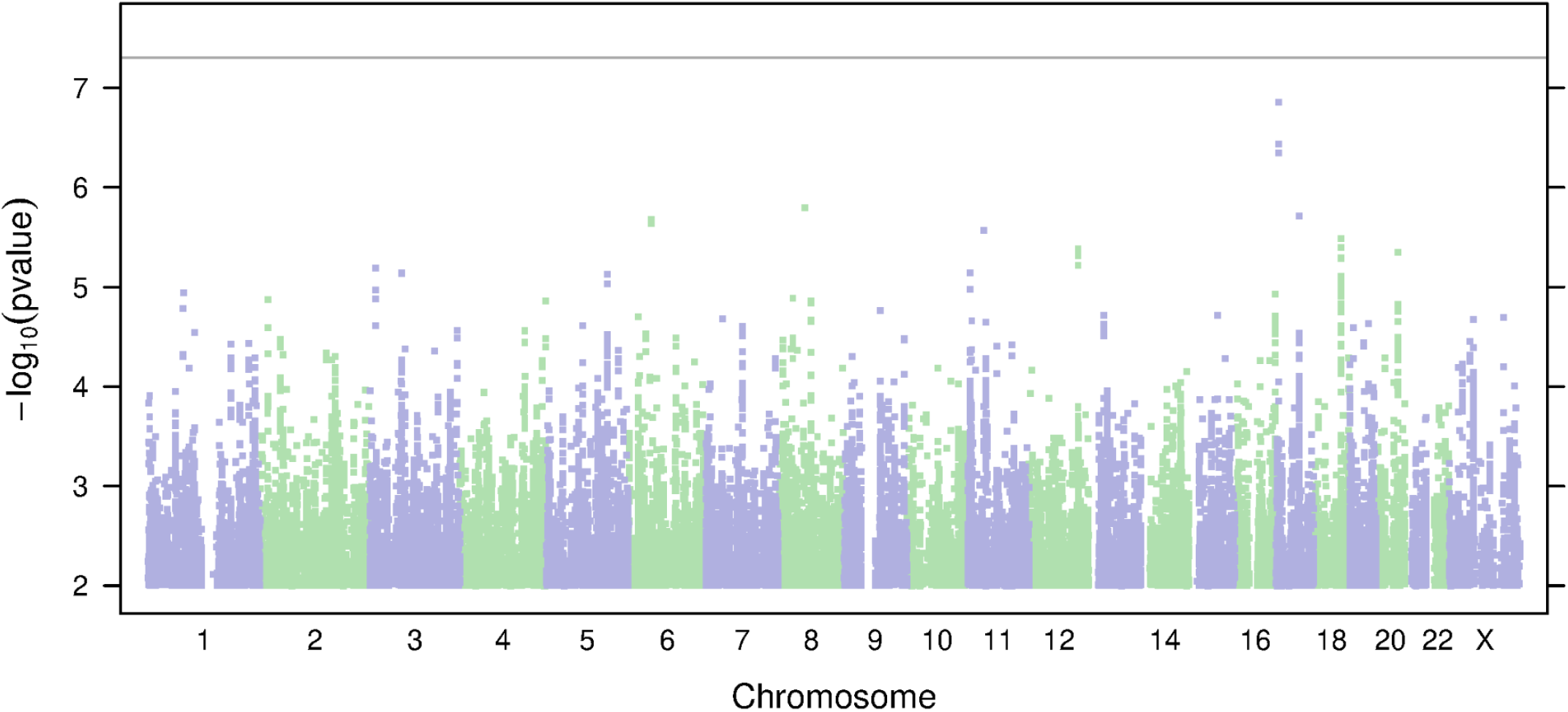
Manhattan plot for the ‘Loss of taste or smell’ phenotype in the South Asian population.

**Supplementary Figure 7:**
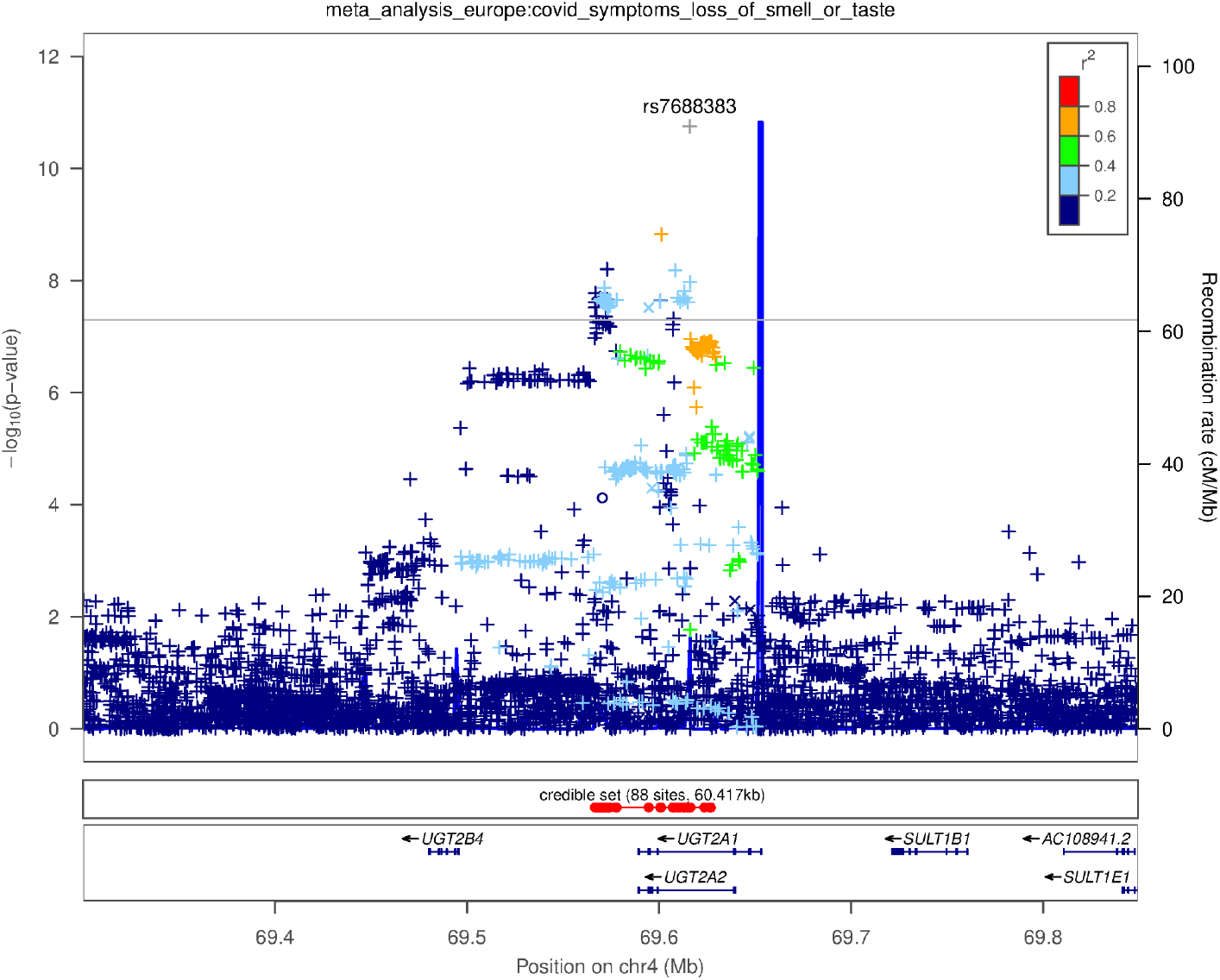
Genetic associations with the ‘Loss of taste or smell’ phenotype. Regional plots around the *UGT2A1* and *UGT2A2* locus in the European population. Colors indicate strength of linkage disequilibrium relative to the index SNP (rs7688383). Imputed variants are indicated with ‘+’ symbols or ‘x’ symbols for coding variants. Where imputed variants weren’t available, directly genotyped variants are indicated by ‘o’ symbols or diamond symbols for coding variants.

**Supplementary Figure 8:**
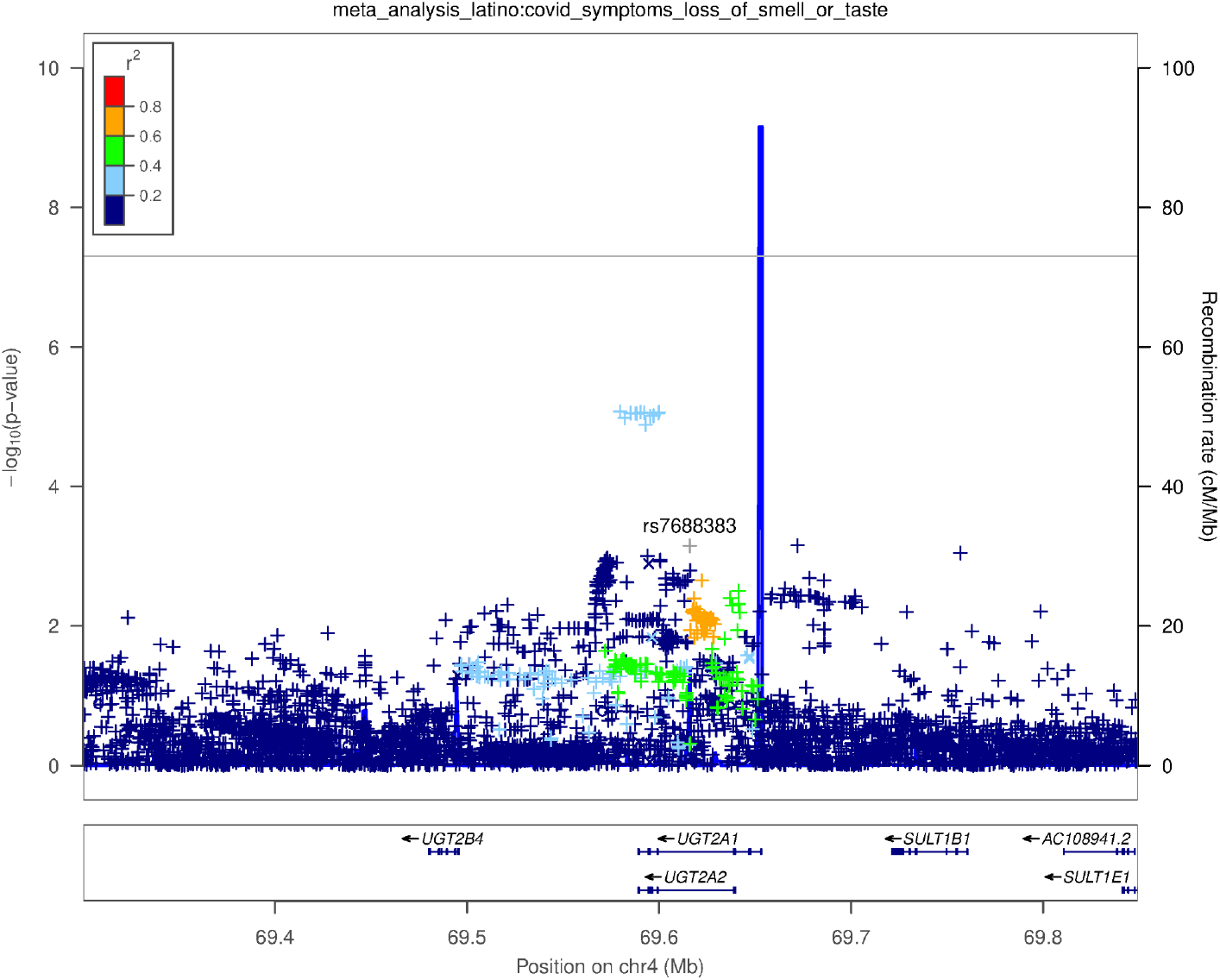
Genetic associations with the ‘Loss of taste or smell’ phenotype. Regional plots around the *UGT2A1*/*UGT2A2* locus in the Latino population. Colors indicate strength of linkage disequilibrium relative to the index SNP (rs7688383). Imputed variants are indicated with ‘+’ symbols or ‘x’ symbols for coding variants. Where imputed variants weren’t available, directly genotyped variants are indicated by ‘o’ symbols or diamond symbols for coding variants.

**Supplementary Figure 9:**
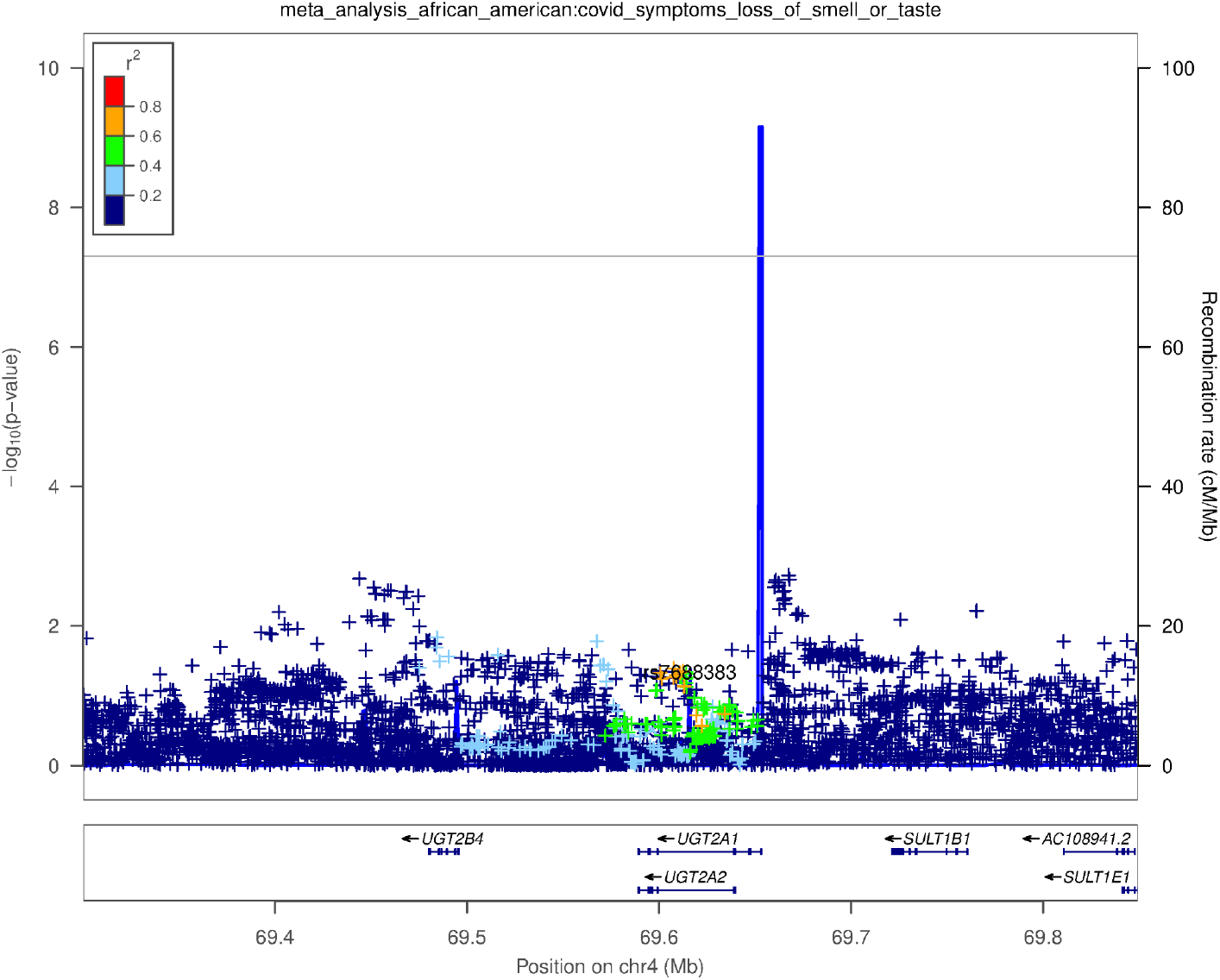
Genetic associations with the ‘Loss of taste or smell’ phenotype. Regional plots around the *UGT2A1*/*UGT2A2* locus in the African American population. Colors indicate strength of linkage disequilibrium relative to the index SNP (rs7688383). Imputed variants are indicated with ‘+’ symbols or ‘x’ symbols for coding variants. Where imputed variants weren’t available, directly genotyped variants are indicated by ‘o’ symbols or diamond symbols for coding variants.

**Supplementary Figure 10:**
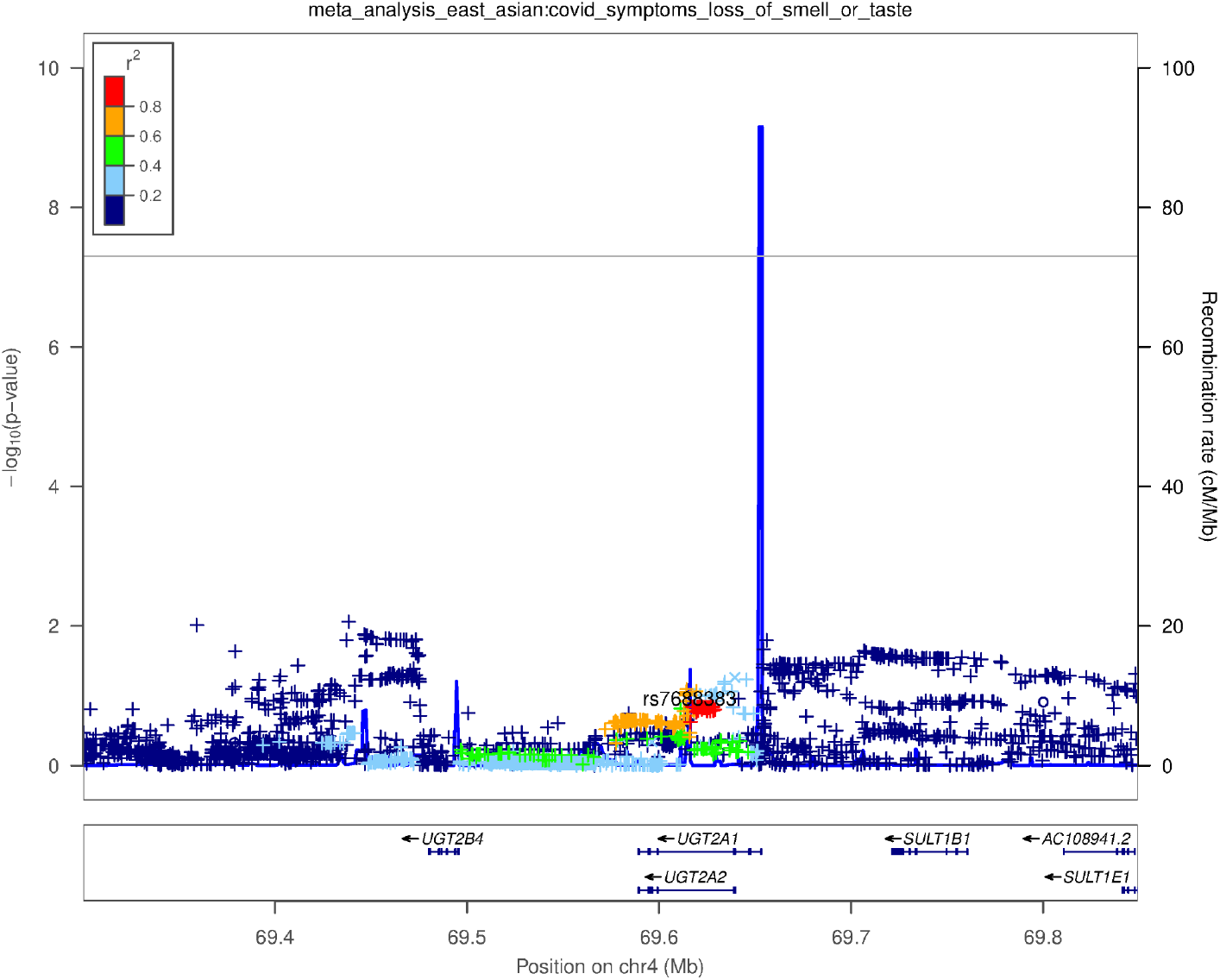
Genetic associations with the ‘Loss of taste or smell’ phenotype. Regional plots around the *UGT2A1*/*UGT2A2* locus in the East Asian population. Colors indicate strength of linkage disequilibrium relative to the index SNP (rs7688383). Imputed variants are indicated with ‘+’ symbols or ‘x’ symbols for coding variants. Where imputed variants weren’t available, directly genotyped variants are indicated by ‘o’ symbols or diamond symbols for coding variants.

**Supplementary Figure 11:**
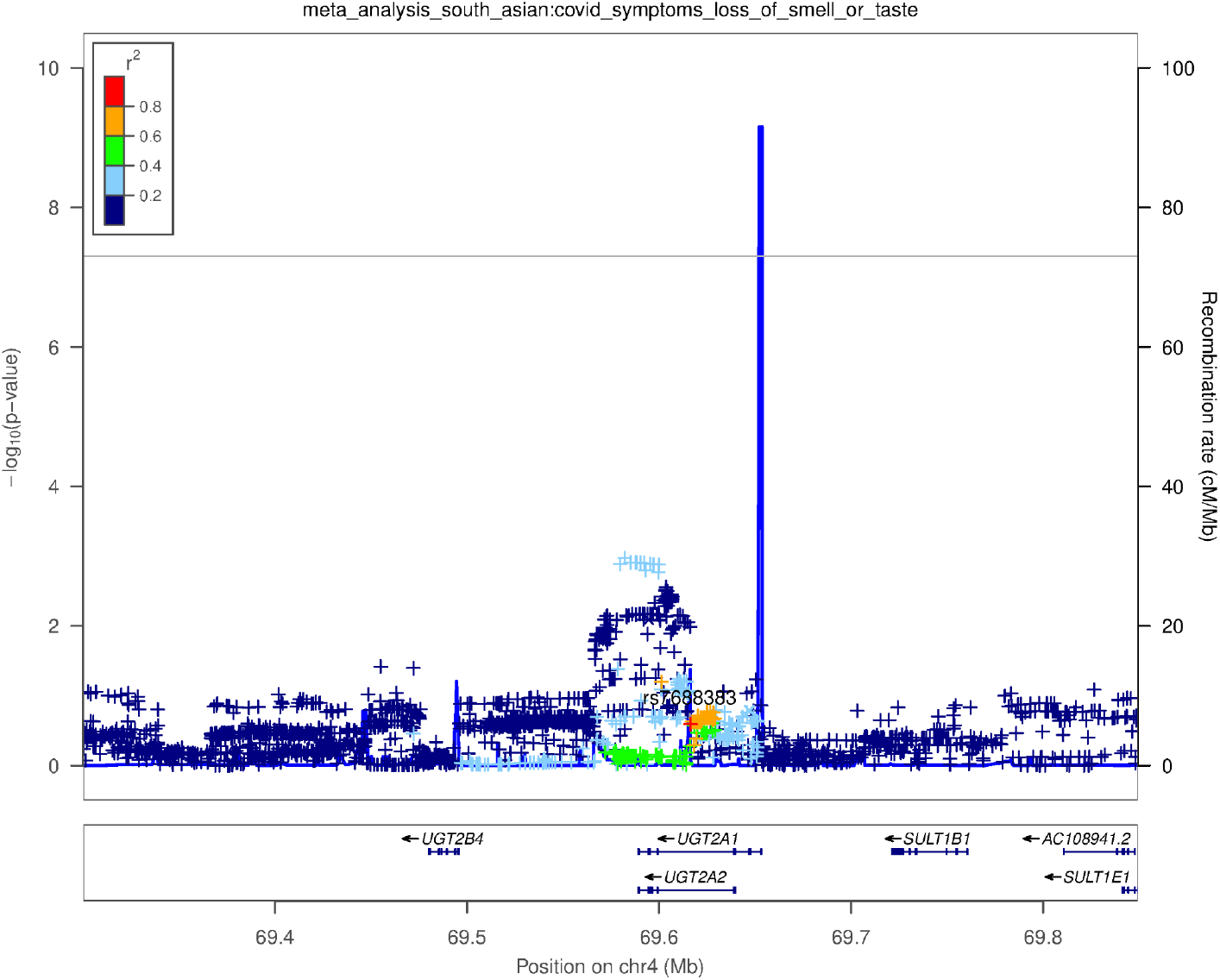
Genetic associations with the ‘Loss of taste or smell’ phenotype. Regional plots around the *UGT2A1*/*UGT2A2* locus in the South Asian population. Colors indicate strength of linkage disequilibrium relative to the index SNP (rs7688383). Imputed variants are indicated with ‘+’ symbols or ‘x’ symbols for coding variants. Where imputed variants weren’t available, directly genotyped variants are indicated by ‘o’ symbols or diamond symbols for coding variants.

